# Calibrated large language models demonstrate a consistent advantage in a randomized longitudinal medical education intervention

**DOI:** 10.64898/2026.10.30.26366878

**Authors:** Jay Khurana, Hossam A. Zaki, Adam Dawer, Ellie Pavlick, Sahil Gupta, Sriya Sai Pushpa Datla, Salma Eldeeb, Saima Chaudhry, Syed A. Rizvi

## Abstract

Medical trainees increasingly use large language models to review lecture material, but the educational value of that output depends on how it is generated, with prompt and model choice rarely measured. We tested a calibrated pipeline converting Neurology Grand Rounds recordings into summaries and quizzes in a year-long randomized study. In Phase 1, six summary and six question prompts across two models were scored with 2,692 rubric annotations. Claude Prompt 0 produced the best summaries (composite 76.9%) and GPT Prompt 1 the best questions (79.6%); quality varied widely by prompt, and neither model won both tasks. In Phase 2, students in 11 clerkship cohorts over a year were randomized within cohort to receive a summary before each quiz. Summary recipients reported reviewing more and studying faster and scored higher in 9 of 11 cohorts (p = 0.026), although the between-student difference was small (+0.28 of 10 points; P = 0.28).

## Introduction

Medical trainees are increasingly turning to large language models (LLMs) to compress, retrieve, and review the growing volume of clinical content they must master.^1,2^ Survey data suggest that generative AI tools are already a routine part of many medical students’ study habits.^3–5^ In neurology in particular, a discipline with dense, hierarchical knowledge structures and a high cognitive load,^6^ Grand Rounds remain a cornerstone of clerkship and residency education, but the volume and pace of content, alongside co-occurring clinical responsibilities, can outstrip trainees’ capacity to consolidate it before clinical duties resume. Grand rounds are often delivered as slide-driven, largely passive presentations with limited opportunity for questions,^7^ and attendees usually leave without companion learning material.

Despite this rapid adoption, much of the published evidence base still consists of studies that benchmark LLM performance against standardized examinations. ChatGPT performed near the U.S. Medical Licensing Examination (USMLE) passing threshold in early evaluations,^8,9^ and on neurology board-style examinations it performed within the range of human test-takers,^10^ with GPT-4 exceeding the mean human score.^11^ Benchmark performance has risen quickly since,^12–14^ which makes the ceiling on this class of question a moving target rather than a finding. These studies remain valuable as feasibility demonstrations, but they evaluate model capability rather than true curricular integration.

Test-enhanced learning, the well-established cognitive-psychology principle that retrieval practice with feedback durably improves retention,^15–20^ provides a strong theoretical motivation for using LLMs to scale weekly quizzing in clerkship education, but only if the generated content is correct, relevant, and aligned with learning objectives. A lecture-specific summary supplied before assessment functions as an advance organizer, giving learners a structure into which new material can be assimilated.^21^ Advance organizers produce small but consistent gains in learning and retention across a large meta-analytic literature,^22^ and supplying that structure externally reduces the extraneous cognitive load of reconstructing it from a lengthy transcript.^23^ This is distinct from summarization carried out by the learner, which reviews of study techniques rate as low utility.^19^

Reviews and early curricular reports of LLMs in pedagogy consistently emphasize that giving learners raw access to a chatbot is not the same as deploying a curated AI pipeline.^24–28^ For the subset of studies where LLMs are applied in a structured manner, the design of the LLM intervention, meaning prompt phrasing, model choice, content structure, and hallucination handling, is rarely treated as a measured variable, even though it is plausibly a dominant determinant of educational effect. In a curricular setting, an educational intervention therefore encompasses not only the underlying model, but also the source material, prompts, and output structure.

This study addresses two gaps. First, prompt and model selection are rarely evaluated against standardized rubrics that capture the dimensions clinicians actually care about: hallucination,^29,30^ learning-objective coverage, and expert-rated clinical relevance.^31–34^ Prompt phrasing is recognized as a clinical competency in its own right,^35^ LLM summaries of medical evidence can be factually inconsistent with their source,^36^ and adaptation strategy determines whether model output matches expert-written text on summarization tasks.^37^ In medical education, this quality control is particularly important because inaccurate or clinically irrelevant generated content may be incorporated into learners’ knowledge. Second, almost no work has prospectively deployed an LLM pipeline as a recurring weekly curricular intervention across multiple cohorts within a single specialty; the closest precedents are tool descriptions and content-generation pipelines validated without learner deployment,^38,39^ or deployments confined to a single cohort.^40^

To address the gap of standardization of intervention quality, we developed a calibrated end-to-end pipeline (Fig. 1a) that ingests recorded Neurology Grand Rounds, transcribes them,^41^ and generates lecture summaries and 10-item MCQ quizzes. We calibrated the pipeline (Phase 1) by scoring six summary prompt variants and six MCQ prompt variants across two LLMs on three full lectures, yielding 2,692 rubric annotations. To address the gap in longitudinal tracking, we then deployed the resulting optimized pipeline (Phase 2) across 11 cohorts of third-year clerkship students under a within-cohort randomized design, comparing students who received AI summaries to those who took the same AI-generated quiz without a summary. We hypothesized that students who received the AI-generated summaries would outperform those who didn’t on the weekly quizzes, while also endorsing improved efficiency and reviewability.

**Fig. 1.**
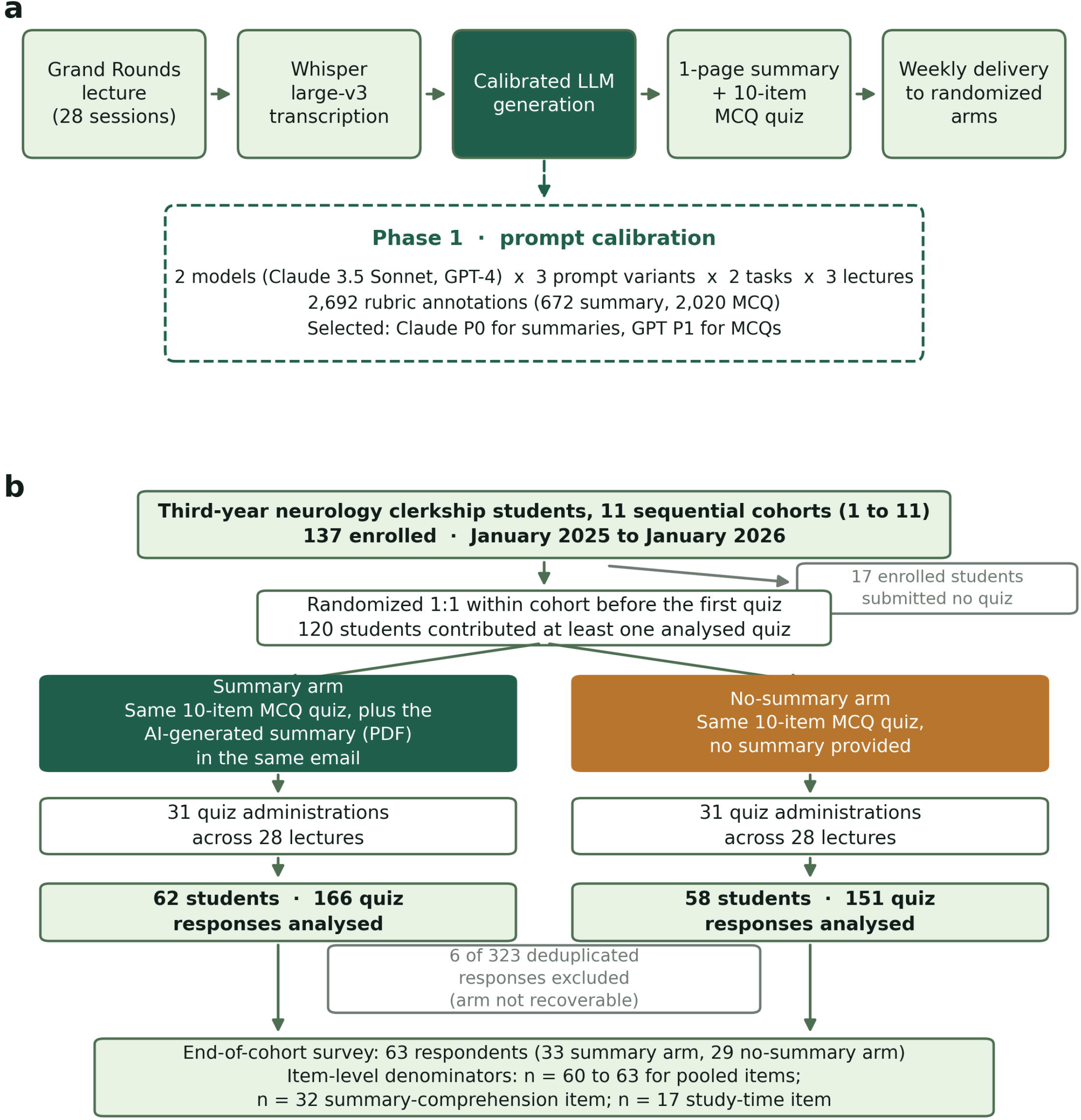
Pipeline architecture and participant flow. a, The four-stage generation pipeline. Recorded Grand Rounds audio is transcribed with Whisper large-v3, processed by a calibrated LLM under a structured prompt to produce a one-page summary and a ten-item MCQ quiz, and delivered to randomized arms. The dashed inset summarizes Phase 1 prompt calibration, in which two models and three prompt variants were scored across two tasks and three lectures using 2,692 rubric annotations, selecting Claude Prompt 0 for summaries and GPT Prompt 1 for MCQs. b, CONSORT-style flow for the Phase 2 randomized intervention. Of 137 students enrolled across 11 sequential cohorts, 17 submitted no quiz and 120 contributed at least one analyzed response (62 Summary, 58 No-summary), giving 317 analyzed responses from 323 collected.

## Results

### Phase 1: Prompt calibration

Composite scores by prompt, computed from the rubric in Table 1, are shown in Fig. 2 and Table 2. Among summary prompts (Fig. 2a), Claude Prompt 0 (P0), chain of thought, achieved the highest composite (76.9%), driven by a 100% hallucination-free rate and the highest learning-objective coverage (82.6%); GPT prompts P1 and P2 hallucinated content on 50% of evaluated lectures, disqualifying them for clinical educational use. Among MCQ prompts (Fig. 2b), GPT Prompt 1 (P1) achieved the highest composite (79.6%), with the highest summary-only answerability (53.8%), the joint-highest hallucination-free rate (100%), and the second-highest expert-rated relevance (85.0%). Notably, no single model dominated both tasks: Claude excelled at summaries, GPT at question generation. Furthermore, the chain-of-thought prompt (P0) outperformed the baseline prompt (P1) for summary generation in both models. The deployed pipeline therefore combined Claude P0 for summaries and GPT P1 for MCQs.

**Table 1.**
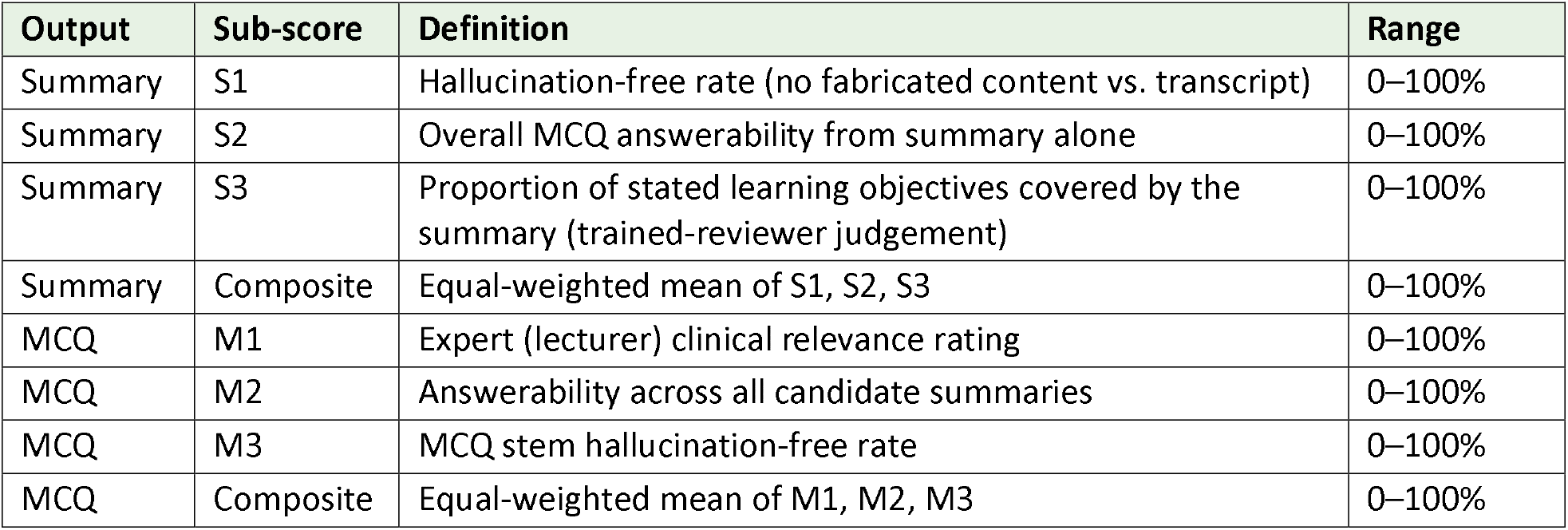
Calibration rubric and composite scoring schema (Phase 1).

**Table 2.**
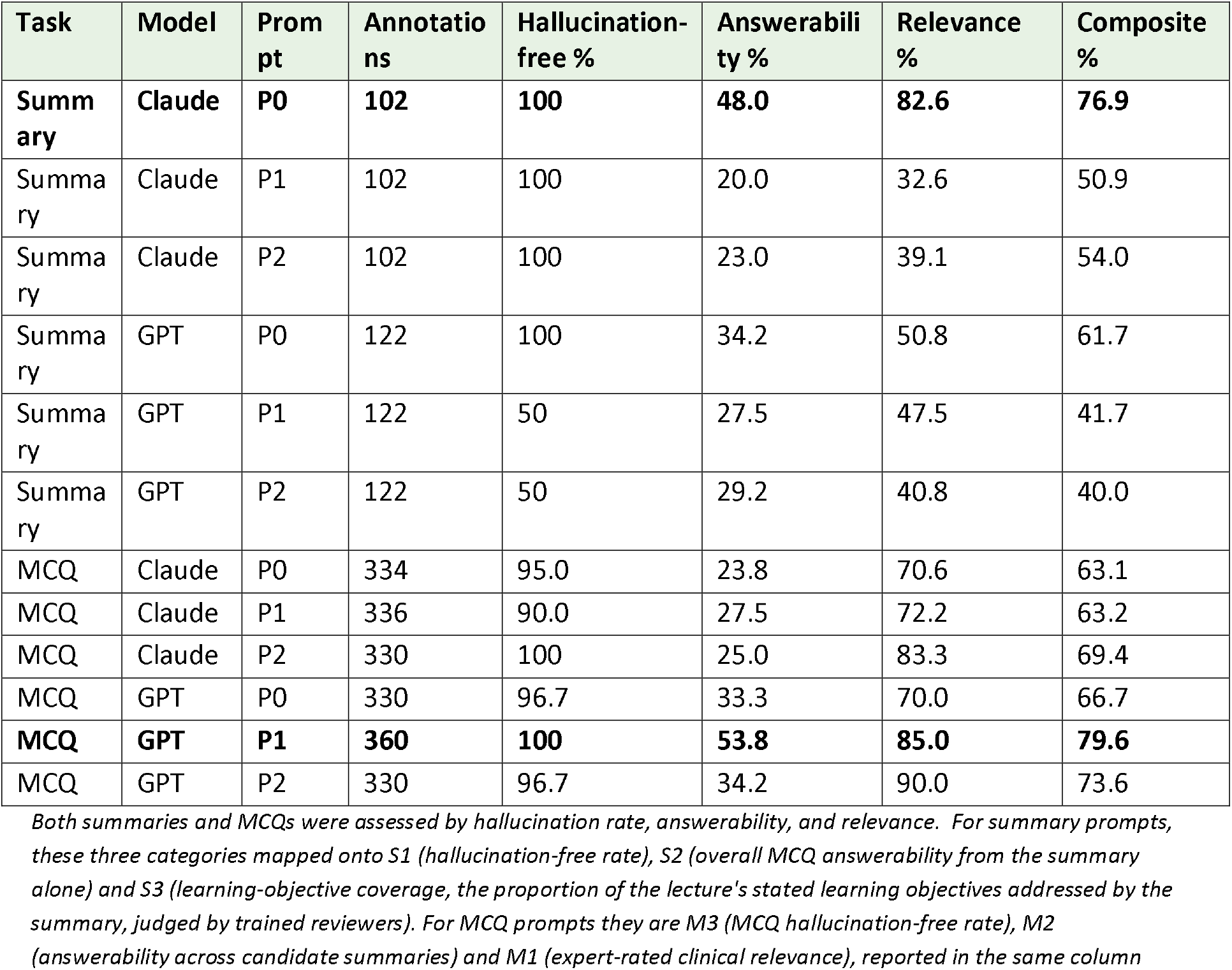
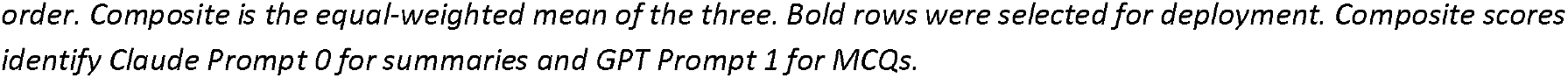
Phase 1 prompt calibration results.

| Task | Model | Prompt | Annotations | Hallucination-free % | Answerability % | Relevance % | Composite % |
| --- | --- | --- | --- | --- | --- | --- | --- |
| **Summary** | **Claude** | **P0** | **102** | **100** | **48.0** | **82.6** | **76.9** |
| Summary | Claude | P1 | 102 | 100 | 20.0 | 32.6 | 50.9 |
| Summary | Claude | P2 | 102 | 100 | 23.0 | 39.1 | 54.0 |
| Summary | GPT | P0 | 122 | 100 | 34.2 | 50.8 | 61.7 |
| Summary | GPT | P1 | 122 | 50 | 27.5 | 47.5 | 41.7 |
| Summary | GPT | P2 | 122 | 50 | 29.2 | 40.8 | 40.0 |
| MCQ | Claude | P0 | 334 | 95.0 | 23.8 | 70.6 | 63.1 |
| MCQ | Claude | P1 | 336 | 90.0 | 27.5 | 72.2 | 63.2 |
| MCQ | Claude | P2 | 330 | 100 | 25.0 | 83.3 | 69.4 |
| MCQ | GPT | P0 | 330 | 96.7 | 33.3 | 70.0 | 66.7 |
| **MCQ** | **GPT** | **P1** | **360** | **100** | **53.8** | **85.0** | **79.6** |
| MCQ | GPT | P2 | 330 | 96.7 | 34.2 | 90.0 | 73.6 |

*Both summaries and MCQs were assessed by hallucination rate, answerability, and relevance. For summary prompts, these three categories mapped onto S1 (hallucination-free rate), S2 (overall MCQ answerability from the summary alone) and S3 (learning-objective coverage, the proportion of the lecture's stated learning objectives addressed by the summary, judged by trained reviewers). For MCQ prompts they are M3 (MCQ hallucination-free rate), M2 (answerability across candidate summaries) and M1 (expert-rated clinical relevance), reported in the same column*
*order. Composite is the equal-weighted mean of the three. Bold rows were selected for deployment. Composite scores identify Claude Prompt 0 for summaries and GPT Prompt 1 for MCQs.*

**Fig. 2.**
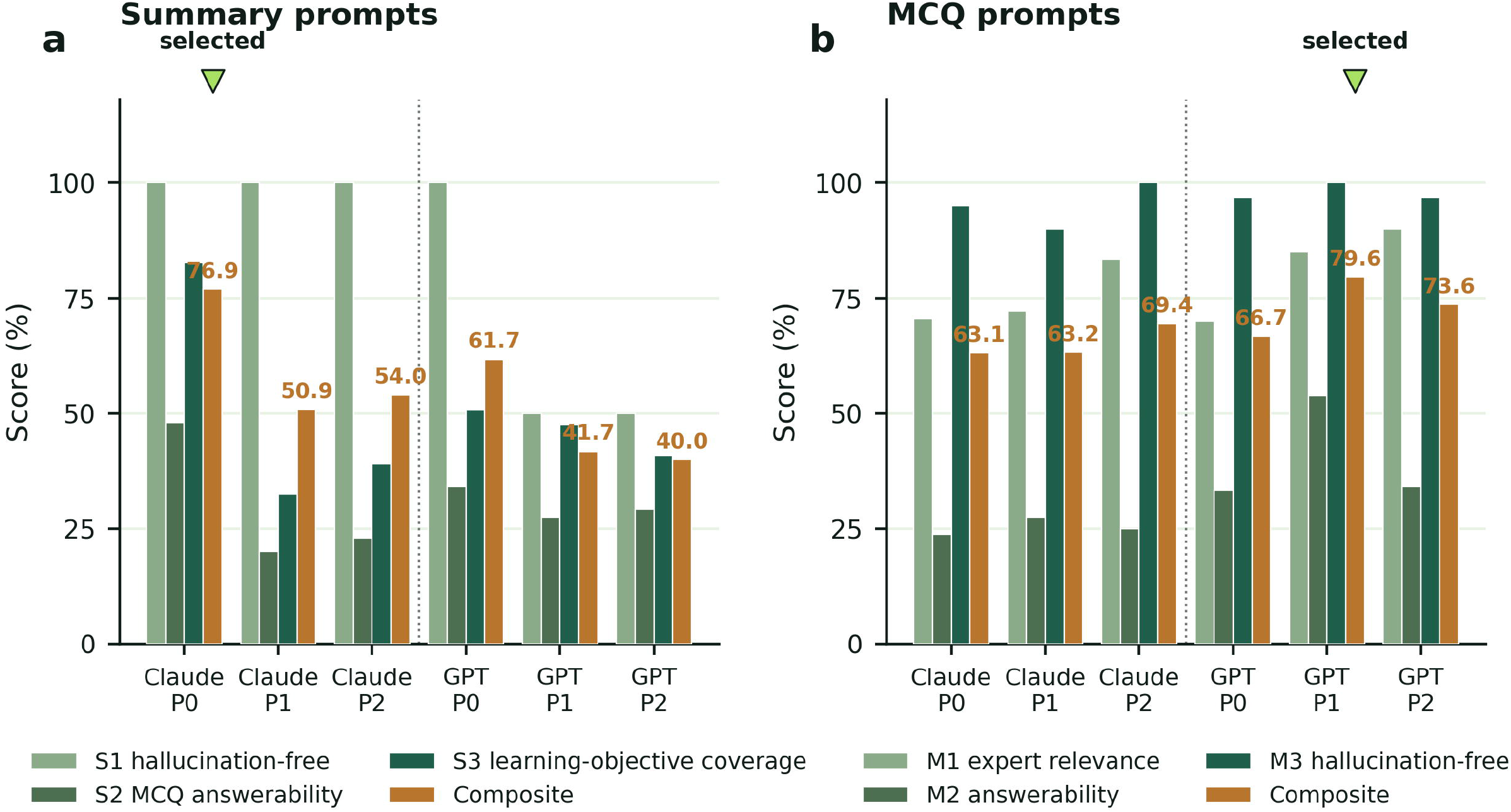
Phase 1 prompt calibration composite scores. a, Summary prompt variants scored on S1 (hallucination-free rate), S2 (overall MCQ answerability from the summary alone) and S3 (learning-objective coverage, the proportion of the lecture’s stated learning objectives addressed by the summary, judged by trained reviewers), with the equal-weighted composite in amber. b, MCQ prompt variants scored on M1 (expert clinical relevance), M2 (answerability across candidate summaries) and M3 (MCQ hallucination-free rate). Green markers indicate the variant selected for deployment.

### Phase 2: Quiz performance

A total of 323 deduplicated quiz responses were collected across 31 quiz administrations(Fig. 1b). Six responses could not be matched to a randomization assignment and were excluded, leaving 317 analyzed responses from 120 unique students, 62 in the Summary arm and 58 in the No-summary arm (Summary arm = 166 responses; No-summary arm = 151 responses).

In the between-student comparison, mean scores were 8.14 in the Summary arm and 7.86 in the No-summary arm, a difference of +0.28 points on a 10-point quiz (95% CI ™0.23 to +0.80; Cohen’s d = +0.20, 95% CI −0.16 to +0.56; P = 0.28). The pooled comparison of all 317 responses gave a similar estimate (+0.29 points, 95% CI −0.11 to +0.69; d = +0.16; P = 0.15) but overstates precision: the intraclass correlation for student was 0.38, implying an effective sample size of approximately 195 rather than 317. A mixed-effects model using all responses returned essentially the same estimate (+0.27 points, 95% CI −0.24 to +0.79; P = 0.30). The distribution of responses in each arm is shown in Fig. 3a; scores in both arms are concentrated in the upper part of the scale, with 65.6% at 8 or above.

**Fig. 3.**
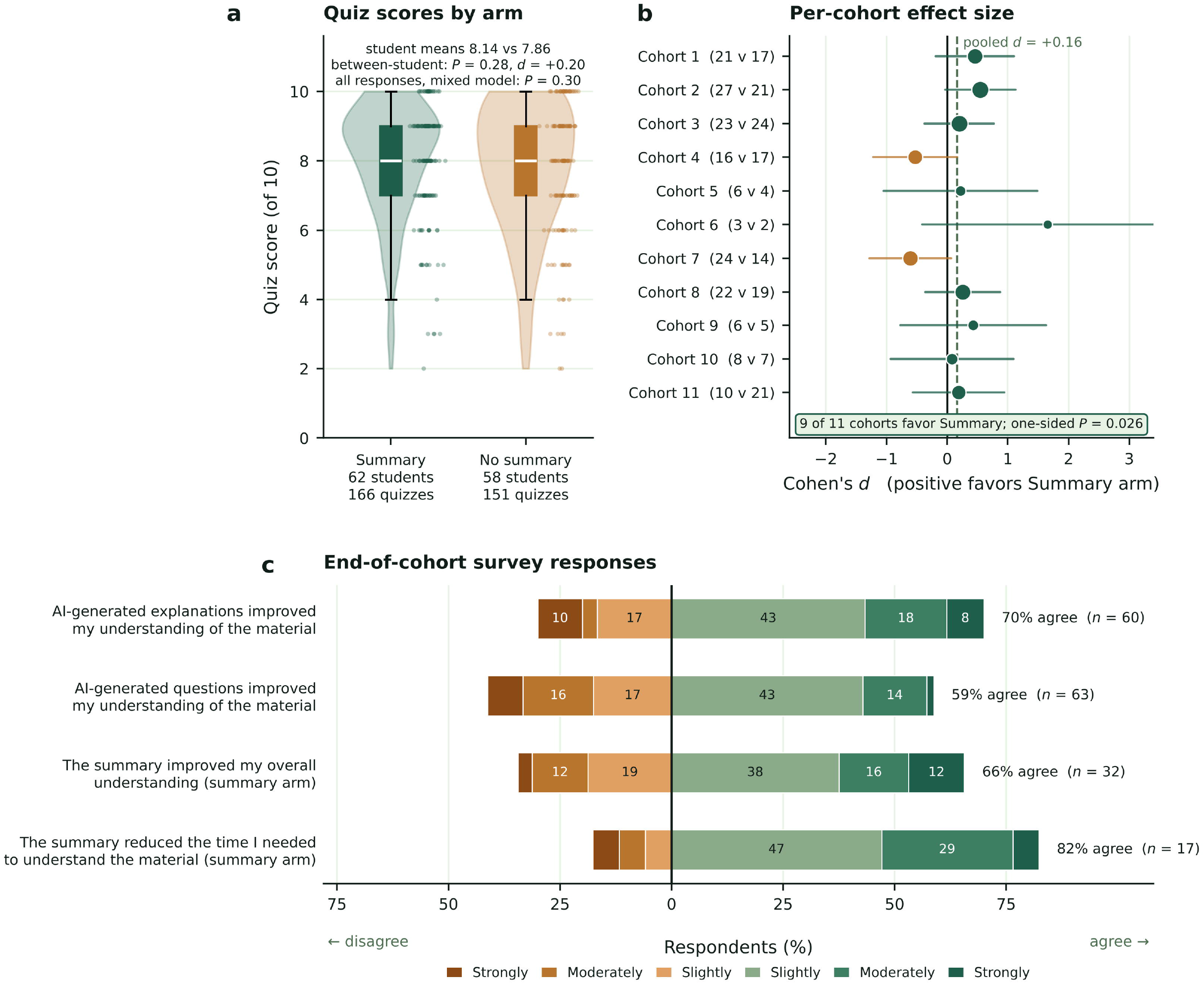
Phase 2 intervention outcomes. a, Distribution of individual quiz scores by arm (Annotations give the principal between-student comparison, in which each student contributes one mean score (Welch’s t-test, P = 0.28; d = +0.20), and the mixed-effects model fitted to all responses (P = 0.30). b, Per-cohort standardized mean difference (Cohen’s d) with approximate 95% confidence intervals; marker area is proportional to total responses, and per-cohort arm sizes are given in parentheses. Cohorts are numbered 1 to 11 in chronological order, and the dashed line marks the pooled response-level d (+0.16). The Summary arm scored higher in 9 of 11 cohorts (randomization test on the number of cohorts favoring Summary, permuting arm assignment among students within cohort, one-sided P = 0.026; exact binomial P = 0.033). c, End-of-cohort survey responses on a six-point Likert scale; denominators differ by item because summary-specific items were shown only to students who reported receiving a summary.

Per-cohort effect sizes are displayed in Fig. 3b, and per-quiz means by arm in Supplementary Fig. S1. The Summary arm scored higher in 9 of 11 cohorts, a pattern unlikely to arise by chance under the randomization test (one-sided P = 0.026, two-sided P = 0.053; Supplementary Fig. S2); the exact binomial sign test gave one-sided P = 0.033 (two-sided P = 0.065), so both two-sided values sit just above 0.05. The two cohorts favoring the No-summary arm were not the smallest: they contributed 33 and 38 responses, more than most cohorts favoring the Summary arm, and their differences (−1.13 and −0.79 points) were among the largest in absolute magnitude. Weighting each cohort’s difference by its number of responses, rather than counting direction alone, gave a smaller advantage that did not reach significance (+0.22 points; one-sided P = 0.21).

### Phase 2: Subjective learning effects

Sixty-three students returned the end-of-cohort survey (Fig. 3c). Item denominators differ because students who reported not having received a summary were not asked the summary-specific items. Among summary recipients, 78.8% (26/33) reported being more likely to review Grand Rounds content as a result of receiving the summary, 82.4% (14/17) reported reduced time required to understand the material, and 65.6% (21/32) reported better overall understanding. 58.7% (37/63) of students reported the questions improved their understanding of Grand Rounds content, with 70% (42/60) agreeing the same of the associated question explanations. The response scale offered no neutral option, so every respondent fell on either the agree or the disagree side of each item, and a substantial majority chose agree on all four questions.

## Discussion

This study provides three concrete contributions to the literature on LLMs in medical education. First, we show that prompt and model selection materially affect educational content quality, and that no single model dominates: Claude P0, the chain-of-thought prompt, produced the cleanest summaries, while GPT P1 produced the most relevant and answerable MCQs. Second, when the calibrated pipeline was deployed in a real curricular setting, students who received AI summaries scored higher than those who did not in 9 of 11 cohorts, a consistent advantage under a randomization test on the direction of the cohort differences (one-sided P = 0.026) that is unaffected by the clustering of repeated quizzes within students, because students are nested within cohorts. Replication of direction across 11 consecutive rotations, with different students, lecturers, and content over 13 months, is the most robust feature of the data. What the study does not establish is magnitude: the between-student difference (+0.28 points, 95% CI −0.23 to +0.80) is small, and weighting cohorts by size rather than counting direction does not reach significance. Third, the subjective effect on study behavior was substantial: more than three-quarters of summary recipients reported being more likely to review content and needing less time to understand it.

The prompt-calibration component of this study addresses the first gap identified in the Introduction, one that recent reviews have also flagged.^24,25,42^ The 36-percentage-point composite-score gap between the best summary prompt (Claude P0, 76.9%) and the worst (GPT P2, 40.0%) underscores how much variance is hidden behind generic LLM usage. This degree of prompt-induced variance is incompatible with publication-grade reproducibility unless prompts are reported in detail and evaluated against a standardized rubric.^30,43^ Prompt design, not model identity alone, governed output quality: within a single model, summary composites varied by up to 26 percentage points across prompt variants (Claude P0, 76.9%; Claude P1, 50.9%). Work on the psychometric properties of model-generated items reaches a compatible conclusion: generated MCQs can approach human-authored items on difficulty and discrimination, but only under specific generation conditions.^32–34^ Most informal use of these tools relies on a single unrefined prompt, which is the condition under which that variance is least visible to the learner. Because no model led on both tasks, multi-model architectures should be the default. Hard-coding a single foundation model into a curriculum is a methodological mistake. In our evaluations, two GPT prompt variants had a 50% hallucination rate, which is incompatible with unsupervised educational use regardless of the model’s headline accuracy. Evaluations of LLM-generated pharmacotherapy simulations found the same weakness: clinical accuracy was the domain most often failed, even where instructional design was rated highly.^44^ Programs and medical schools evaluating commercial AI study tools should therefore ask not only which model a tool uses and how it performs on benchmarks, but also how its prompts were calibrated and against what rubric. Emerging competency frameworks for AI in medical education make the same point from the curricular side.^45,46^

At most institutions, Neurology Grand Rounds and other structured educational activities offer no companion learning material, leaving participants with only the knowledge they obtained directly from the lecture. Teaching time is also vulnerable to interruption: among surgical residents, disruptive interruptions were about three times as frequent during feedback, coaching and informal teaching as across the workday overall,^47^ and attending rounds averaged 12.6 interruptions per session.^48^ A long line of work in cognitive psychology shows that retrieval practice, meaning repeated and spaced testing with feedback, produces substantially stronger long-term retention than re-reading the same material.^15,18,19^ Our pipeline is, in effect, a scaled implementation of test-enhanced learning grafted onto Grand Rounds. The direction held in 9 of 11 cohorts (randomization test on cohort direction, one-sided P = 0.026), which is compatible with a small per-encounter benefit from the weekly dose, one quiz per week with the summary supplied alongside it, although the size of that benefit is not resolved at this sample size. Students’ own reports point the same way, with 78.8% of summary recipients reporting being more likely to review Grand Rounds content, 82.4% reporting reduced time required to understand the material, and 65.6% reporting better overall understanding. We would expect that increasing the dose, for example daily micro-quizzes or spaced repetition over the rotation, and pairing each summary with retrieval-practice questions in the summary itself would amplify the effect; this is a focus of ongoing work.

Our findings sit within a widening but still heterogeneous body of interventional evidence. The 50-physician trial of LLM-augmented diagnostic reasoning^49^ found no measurable improvement in physician performance even though the LLM alone outperformed all physician groups, suggesting that the interface between clinician and model, rather than model capability, is the bottleneck. By contrast, the Gan et al. orthopedic-education trial^50^ in 129 medical students found significant improvement in subject-specific examination performance after a two-week intervention, and the Brügge et al. patient-simulation trial^51^ of 21 students reported clinical-reasoning gains after four feedback sessions. A generative teaching assistant deployed for personalized learning,^52^ a digital serious game for medical terminology,^53^ an AI-based OSCE platform used during licensure preparation,^54^ and a three-arm trial of digital versus face-to-face peer support^55^ report gains in engagement or performance, in designs ranging from usage analyses to randomized trials. The pattern across this literature is consistent with our own: when the LLM is delivered as structured, calibrated, prepared content rather than as a chatbot the student must drive, measurable improvements emerge; when it is offered as an open-ended assistant, they often do not.^49^ Our study extends this pattern to a full academic year of weekly delivery across 11 consecutive cohorts.

Several limitations qualify these findings. The study was conducted at a single U.S. academic medical center with third-year clerkship students, so generalization to other training programs and specialties requires replication. Three limitations bear most directly on the strength of the inference. First, the study is underpowered for the individual-level effect: detecting the observed between-student effect (d = 0.20) at 80% power and two-sided α = 0.05 would require roughly 394 students per arm, against the 62 and 58 analyzed. Second, arm assignment was fixed at the student level, so repeated quizzes are clustered (ICC 0.38) and the effective sample is nearer 195 than 317 responses; randomization keeps the point estimate unbiased, but precision is lower than the raw response count suggests. Third, and most consequentially for detecting an effect, the instrument is compressed against its ceiling. Quizzes are marked out of 10, and the distribution is left-skewed (skewness −1.03), with 65.6% of responses at 8 or above, 45.4% at 9 or above, and 17.0% at 10, leaving a mean of only 2.15 unearned items per response in the control arm within which any benefit could appear. Restriction of range attenuates observable differences, and in item-response terms, a test whose items are far easier than the examinees’ ability carries little information about those examinees: items answered correctly by nearly everyone have near-zero discrimination, so the upper half of the ability distribution is measured poorly. The items were also predominantly recall-level, whereas the benefits of retrieval practice are typically largest for higher-order transfer, so the instrument probed the level at which a summary would be expected to help least. Consistent with attenuation, restricting the analysis to the 22 of 31 quiz administrations with a mean below 85% yielded a larger pooled estimate (+0.40 points, d = +0.21), although this remained non-significant (P = 0.11) and is suggestive rather than confirmatory. A more discriminating instrument, with items targeted near 50 to 70% difficulty, a greater number of items, application-level and clinical-vignette formats, and item-response-theory scoring in place of raw sum scores, would materially increase the sensitivity of a replication. Self-report bias is likely in the qualitative survey, since students who received the summary may have over-reported its perceived value, and survey denominators vary by item. Quizzes were administered in relative proximity to the lecture, so we did not test delayed recall, which is the dimension on which test-enhanced learning shows its largest benefit. Finally, newer model variants became available during the study period but were not re-evaluated through the full Phase 1 protocol, so the specific prompt and model rankings reported here should be read as a demonstration of the calibration method rather than as durable claims about particular models.

The natural next steps are a multi-site replication with pre-registered powering for a per-individual effect of d = 0.20; addition of a delayed-recall outcome at 4 to 8 weeks post-lecture to assess the test-enhanced-learning hypothesis directly; re-evaluation of current foundation models under the same Phase 1 rubric to determine whether multi-model dominance has collapsed; inclusion of retrieval-practice questions within the summary itself, converting it from a passive review aid into an active study tool; and a head-to-head comparison of the calibrated pipeline against a generic chatbot-as-tutor condition to isolate the marginal value of calibration.

A systematically calibrated end-to-end LLM pipeline produced educationally valid neurology summaries and MCQs, was feasible to deliver weekly across a full academic year, and produced a consistent cohort-level advantage in quiz performance (9 of 11 cohorts; randomization test, one-sided P = 0.026), with substantial self-reported benefits to review behavior and time to understand. The magnitude of the individual-level effect was small and is not resolved by a study of this size, which a larger, pre-registered trial using a more discriminating instrument should address. Different models excelled at different sub-tasks, supporting a task-specific multi-model architecture. The dominant methodological lesson is that prompt and model selection are first-class educational variables, not implementation details, and the practical lesson, offered as a hypothesis for future testing rather than as a demonstrated result, is that AI in medical education is most likely to help when it is deployed as calibrated curricular content rather than as a chatbot.

## Methods

### Setting and participants

The study was conducted during the third-year neurology clerkship rotation at a single allopathic medical school for the weekly Grand Rounds series from January 2025 through January 2026. One hundred and thirty-seven third-year medical students rotated through 11 sequential cohorts, and 120 unique students contributed at least one analyzed response (Fig. 1b). All Grand Rounds lectures were recorded with speaker permission and processed through the pipeline, and 31 quiz administrations were delivered across the 11 cohorts. Participation was voluntary, and quiz scores were not used for academic grading; completing a quiz counted toward attendance.

### Pipeline architecture

The end-to-end pipeline (Fig. 1) consisted of four stages. Recorded Grand Rounds audio was first transcribed using OpenAI Whisper (large-v3).^41^ Transcripts were then processed by an LLM with a structured prompt to produce both a formatted summary and a 10-item multiple-choice question (MCQ) quiz. Outputs were spot-checked for obvious failures before release, and the resulting summaries and quizzes were distributed to students via email within a week after the lecture.

### Phase 1: Prompt calibration

Three lectures spanning diverse neurology subdomains were used for calibration: Neurogenic Bladder (September 4, 2024), Neurovascular Emergencies (October 16, 2024), and Alternative Therapies in ALS (October 30, 2024). Two LLMs were evaluated: Anthropic’s Claude 3.5 Sonnet and OpenAI’s GPT-4 (June 2024 versions). Three prompt variants per task per model were authored: a chain-of-thought prompt (P0), a baseline prompt (P1), and a learning-objective-anchored prompt (P2), yielding six summary variants and six MCQ variants per lecture.

Each output was scored against a standardized rubric (Table 1). Summaries were scored on three dimensions: (S1) hallucination-free rate, the proportion of tested lectures with no hallucinated content, scored binarily by a trained reviewer with reference to the original transcript; (S2) MCQ answerability, the proportion of accompanying MCQs whose correct answer is derivable from the summary alone; (S3) learning-objective coverage, the proportion of the lecture’s stated learning objectives addressed by the summary, judged by trained reviewers against the lecturer’s objective list. MCQs were scored on three dimensions: (M1) expert relevance, the proportion of MCQs rated clinically relevant by the lecturer, a board-certified neurologist; (M2) answerability, the proportion of MCQs answerable using any of the four candidate summaries; (M3) MCQ hallucination-free rate, the proportion of MCQ stems containing no fabricated content.

Composite scores were computed per output type as the equal-weighted mean of the three sub-scores (33.3% each). The optimal prompt for each task (summary, MCQ) was the one with the highest composite score. Annotations totaled 2,692.

### Phase 2: Randomized intervention

The optimized pipeline was deployed weekly across 31 Grand Rounds lectures spanning 11 cohorts. Within each cohort, students were randomized to one of two arms (Fig. 1b). Students assigned to the Summary arm received the AI-generated lecture summary as a PDF along with the quiz in the same email, while students assigned to the No-summary arm took the same AI-generated quiz with no summary provided. Randomization was performed at the individual level within each cohort before the first quiz. The same 10-item MCQ quiz was delivered to both arms via a Qualtrics or Google Forms link. Each MCQ was scored binarily and totaled out of 10. When a student submitted multiple attempts on the same quiz, only the first submitted score was used.

After the final quiz of each cohort, students received an end-of-cohort qualitative survey (six-point Likert items with no neutral option, plus free-text) capturing self-reported review behavior, perceived comprehension, and time to understand.

### Statistical analysis

The outcome was the per-quiz score (0 to 10). The approved protocol pre-specified a directional hypothesis, that students receiving AI-generated summaries would outperform those who did not.

Because arm assignment was fixed at the student level before the first quiz, the student is the unit of randomization. We therefore take as the principal individual-level analysis a between-student comparison in which each student contributes a single mean quiz score, compared by Welch’s t-test^56^ with Cohen’s d (pooled SD)^57^ as the effect-size measure. This matches the randomization unit and makes no assumption about the correlation among a student’s repeated quizzes. The pooled comparison of all responses is reported alongside it, together with the intraclass correlation for student, because most students contributed more than one quiz (mean 2.6, range 1 to 4) and that comparison assumes an independence the data do not support. A linear mixed-effects model with a random intercept for student, fitted to all responses, is reported as a sensitivity analysis.

At cohort level, because students are nested within cohorts, and no student appears in more than one cohort or arm, cohort-level inference is unaffected by within-student correlation. The exact binomial sign test is reported alongside as the conventional reference; it assumes each cohort is equally likely to favor either arm, which unequal within-cohort allocation makes only approximately true. The confirmatory analysis is a randomization test on the number of cohorts in which the Summary arm scored higher, reproducing the assignment mechanism: arm labels were permuted among students within cohort over 100,000 permutations, preserving each cohort’s allocation ratio and keeping every student’s responses together. The same permutation distribution was used to test the size-weighted mean of the cohort differences, which uses magnitude as well as direction. As the protocol specified a direction, one-sided probabilities are reported, with two-sided values obtained by doubling.

All analyses used Python 3.11 with NumPy,^58^ SciPy,^59^ statsmodels^60^ and Matplotlib.^61^

### Ethics

The study was reviewed by Lifespan Institutional Review Board 2 at Rhode Island Hospital (project 414824, determination 2239881-1, effective 20 October 2024) and received a determination of RESEARCH, NOT HUMAN SUBJECTS RESEARCH. Protocol version 1.0.0 was finalized on 1 October 2024 and specified the directional primary hypothesis, the qualitative secondary endpoints, and a commitment to freeze the generation pipeline before enrollment. Participation was voluntary, quiz performance was not used for academic grading, and no identifiable student data are reported.

### Reporting

The trial is reported in accordance with the CONSORT-AI extension.^62^ Because the intervention was an educational activity determined not to constitute human subjects research, the study was not registered in a clinical trials registry. Items from the TRIPOD-LLM reporting guideline that apply to a deployed generative pipeline are addressed in the Methods.^43^

## Supporting information

Figures with legends, including Supplementary Figures S1 and S2

## Data availability

De-identified quiz response data, per-cohort summary statistics and the Phase 1 rubric annotations are available from the corresponding author on reasonable request.

## Code availability

The analysis code, including the verification script that regenerates every statistic reported here from the underlying response data, is available from the corresponding author on reasonable request.

## Acknowledgements

We thank the Grand Rounds speakers at Brown University Health for permission to record and process their lectures, and the third-year neurology clerkship students who took part. This study received no external funding.

## Author contributions

J.K., H.A.Z., E.P., S.C. and S.A.R. contributed to the conception and methodology of the study, including its design. J.K. and H.A.Z. built the generation pipeline and performed the statistical analysis. J.K., H.A.Z. and A.D. conducted the analyses and drafted the manuscript. H.A.Z. designed the calibration rubric and supervised annotation. S.G., S.S.P.D., and S.E. were responsible for data labeling. S.C. and S.A.R. provided expert neurological review of generated content, facilitated access to the Grand Rounds series and supervised the clerkship integration. All authors interpreted the results, revised the manuscript critically for important intellectual content and approved the final version.

## References

1. Densen, P. Challenges and opportunities facing medical education. Trans. Am. Clin. Climatol. Assoc. 122, 48–58 (2011).

2. Wynter, L., Burgess, A., Kalman, E., Heron, J. E. & Bleasel, J. Medical students: what educational resources are they using? BMC Med. Educ. 19, 36 (2019).

3. Popa, D.-I. et al. Assessing Generative AI Adoption, Tool Preferences, and Cognitive Reliance Among Medical Students: A Cross-Sectional Study. Int. Med. Educ. 5, 66 (2026).

4. Abouammoh, N. et al. Perceptions and Earliest Experiences of Medical Students and Faculty With ChatGPT in Medical Education: Qualitative Study. JMIR Med. Educ. 11, e63400 (2025).

5. Fu, L. et al. Mixed-methods study on GenAI Usage, dependence behaviors, and standardized application paths among Chinese medical students. npj Digit. Med. 9, 714 (2026).

6. Zinchuk, A. V., Flanagan, E. P., Tubridy, N. J., Miller, W. A. & McCullough, L. D. Attitudes of US medical trainees towards neurology education: ‘Neurophobia’ - a global issue. BMC Med. Educ. 10, 49 (2010).

7. Van Hoof, T. J., Monson, R. J., Majdalany, G. T., Giannotti, T. E. & Meehan, T. P. A Case Study of Medical Grand Rounds: Are We Using Effective Methods? Acad. Med. 84, 1144–1151 (2009).

8. Kung, T. H. et al. Performance of ChatGPT on USMLE: Potential for AI-assisted medical education using large language models. PLOS Digit. Health 2, e0000198 (2023).

9. Gilson, A. et al. How Does ChatGPT Perform on the United States Medical Licensing Examination (USMLE)? The Implications of Large Language Models for Medical Education and Knowledge Assessment. JMIR Med. Educ. 9, e45312 (2023).

10. Chen, T. C. et al. Assessment of ChatGPT’s performance on neurology written board examination questions. BMJ Neurol. Open 5, e000530 (2023).

11. Schubert, M. C., Wick, W. & Venkataramani, V. Performance of Large Language Models on a Neurology Board–Style Examination. JAMA Netw. Open 6, e2346721 (2023).

12. Singhal, K. et al. Large language models encode clinical knowledge. Nature 620, 172–180 (2023).

13. Newton, P. M. et al. Can ChatGPT-4o Really Pass Medical Science Exams? A Pragmatic Analysis Using Novel Questions. Med. Sci. Educ. 35, 721–729 (2025).

14. Çelik, A., Kırış, T., Kocabaş, U., Özdemir, E. & Karaca, M. Psychometric characterization of human and artificial intelligence performance on cardiology residency in-service examination items. npj Digit. Med. https://doi.org/10.1038/s41746-026-03002-9 (2026) doi:10.1038/s41746-026-03002-9.

15. Roediger, H. L. & Karpicke, J. D. Test-Enhanced Learning: Taking Memory Tests Improves Long-Term Retention. Psychol. Sci. 17, 249–255 (2006).

16. Larsen, D. P., Butler, A. C. & Roediger III, H. L. Test-enhanced learning in medical education. Med. Educ. 42, 959–966 (2008).

17. Karpicke, J. D. & Blunt, J. R. Retrieval Practice Produces More Learning than Elaborative Studying with Concept Mapping. Science 331, 772–775 (2011).

18. Larsen, D. P., Butler, A. C. & Roediger III, H. L. Comparative effects of test-enhanced learning and self-explanation on long-term retention. Med. Educ. 47, 674–682 (2013).

19. Dunlosky, J., Rawson, K. A., Marsh, E. J., Nathan, M. J. & Willingham, D. T. Improving Students’ Learning With Effective Learning Techniques: Promising Directions From Cognitive and Educational Psychology. Psychol. Sci. Public Interest 14, 4–58 (2013).

20. Green, M. L., Moeller, J. J. & Spak, J. M. Test-enhanced learning in health professions education: A systematic review: BEME Guide No. 48. Med. Teach. 40, 337–350 (2018).

21. Ausubel, D. P. The use of advance organizers in the learning and retention of meaningful verbal material. J. Educ. Psychol. 51, 267–272 (1960).

22. Luiten, J., Ames, W. & Ackerson, G. A Meta-analysis of the Effects of Advance Organizers on Learning and Retention. Am. Educ. Res. J. 17, 211–218 (1980).

23. Young, J. Q., Van Merrienboer, J., Durning, S. & Ten Cate, O. Cognitive Load Theory: Implications for medical education: AMEE Guide No. 86. Med. Teach. 36, 371–384 (2014).

24. Lucas, H. C., Upperman, J. S. & Robinson, J. R. A systematic review of large language models and their implications in medical education. Med. Educ. 58, 1276–1285 (2024).

25. Preiksaitis, C. & Rose, C. Opportunities, Challenges, and Future Directions of Generative Artificial Intelligence in Medical Education: Scoping Review. JMIR Med. Educ. 9, e48785 (2023).

26. Abd-alrazaq, A. et al. Large Language Models in Medical Education: Opportunities, Challenges, and Future Directions. JMIR Med. Educ. 9, e48291 (2023).

27. Divito, C. B., Katchikian, B. M., Gruenwald, J. E. & Burgoon, J. M. The tools of the future are the challenges of today: The use of ChatGPT in problem-based learning medical education. Med. Teach. 46, 320–322 (2024).

28. Magalhães Araujo, S. & Cruz-Correia, R. Incorporating ChatGPT in Medical Informatics Education: Mixed Methods Study on Student Perceptions and Experiential Integration Proposals. JMIR Med. Educ. 10, e51151 (2024).

29. Ji, Z. et al. Survey of Hallucination in Natural Language Generation. ACM Comput. Surv. 55, 248 (2023).

30. Tam, T. Y. C. et al. A framework for human evaluation of large language models in healthcare derived from literature review. npj Digit. Med. 7, 258 (2024).

31. Kiyak, Y. S. & Emekli, E. ChatGPT prompts for generating multiple-choice questions in medical education and evidence on their validity: a literature review. Postgrad. Med. J. 100, 858–865 (2024).

32. Kiyak, Y. S., Kaya, A. B. & Emekli, E. Validity of AI-generated multiple-choice questions in medical education: a systematic review. Postgrad. Med. J. https://doi.org/10.1093/postmj/qgag057 (2026) doi:10.1093/postmj/qgag057.

33. Linde, P. et al. Psychometric properties and detectability of GPT-4o–generated multiple-choice questions compared with human-authored items across imaging specialties. npj Digit. Med. 9, 132 (2026).

34. Qian, W. et al. Performance of DeepSeek in the generation of in-training examination questions in radiology resident education. npj Digit. Med. 9, 384 (2026).

35. Meskó, B. Prompt Engineering as an Important Emerging Skill for Medical Professionals: Tutorial. J. Med. Internet Res. 25, e50638 (2023).

36. Tang, L. et al. Evaluating large language models on medical evidence summarization. npj Digit. Med. 6, 158 (2023).

37. Van Veen, D. et al. Adapted large language models can outperform medical experts in clinical text summarization. Nat. Med. 30, 1134–1142 (2024).

38. Nam, Y. et al. Human-in-the-loop validation of a sequential multi-LLM medical education pipeline. npj Digit. Med. https://doi.org/10.1038/s41746-026-02978-8 (2026) doi:10.1038/s41746-026-02978-8.

39. Pendergrast, T. & Chalmers, Z. Anki Tagger: A Generative AI Tool for Aligning Third-Party Resources to Preclinical Curriculum. JMIR Med. Educ. 9, e48780 (2023).

40. Khurana, J. et al. From lectures to learning outcomes: meaningful integration of AI-generated content in pre-clerkship medical training. BMC Med. Educ. https://doi.org/10.1186/s12909-026-09975-1 (2026) doi:10.1186/s12909-026-09975-1.

41. Radford, A. et al. Robust speech recognition via large-scale weak supervision. in Proc. 40th International Conference on Machine Learning vol. 202 28492–28518 (PMLR, 2023).

42. Jung, E., Durning, S. J. & Schuwirth, L. Health professions education (HPE) in the age of generative AI. npj Digit. Med. https://doi.org/10.1038/s41746-026-03042-1 (2026) doi:10.1038/s41746-026-03042-1.

43. Gallifant, J. et al. The TRIPOD-LLM reporting guideline for studies using large language models. Nat. Med. 31, 60–69 (2025).

44. Farrag, A. N., El-Zeiny, A. & Ali, A. M. Evaluating large language models for pharmacotherapy simulations: a mixed-methods study. npj Digit. Med. 9, 355 (2026).

45. Hunt, V. M. et al. What the AI era doctor should know: a scoping review of proposed artificial intelligence competencies for medical education. npj Digit. Med. 9, 732 (2026).

46. McGrath, S. P., Kim, K. K., Johl, K., Wang, H. & Anderson, N. AI-PACE: a framework for integrating AI into medical education. npj Digit. Med. 9, 595 (2026).

47. Zhang, C. J., Walser, E., Mierzwa, A. & Ott, M. Learning Interrupted: Educational Impact of Interruptions on Surgical Residents. J. Surg. Educ. 79, 875–884 (2022).

48. Armendariz, J. et al. Interruptions to Attending Physician Rounds and Their Effect on Resident Education. J. Grad. Med. Educ. 13, 266–275 (2021).

49. Goh, E. et al. Large Language Model Influence on Diagnostic Reasoning: A Randomized Clinical Trial. JAMA Netw. Open 7, e2440969 (2024).

50. Gan, W. et al. Integrating ChatGPT in Orthopedic Education for Medical Undergraduates: Randomized Controlled Trial. J. Med. Internet Res. 26, e57037 (2024).

51. Brügge, E. et al. Large language models improve clinical decision making of medical students through patient simulation and structured feedback: a randomized controlled trial. BMC Med. Educ. 24, 1391 (2024).

52. Thesen, T. & Park, S. H. A generative AI teaching assistant for personalized learning in medical education. npj Digit. Med. 8, 627 (2025).

53. Kimiafar, K. et al. Evaluating a digital serious game for learning medical terminology in a randomized controlled trial. npj Digit. Med. 9, 427 (2026).

54. Song, J. W. et al. Naturalistic adoption and deliberate practice use of an AI-based OSCE platform during national licensure preparation. npj Digit. Med. https://doi.org/10.1038/s41746-026-03024-3 (2026) doi:10.1038/s41746-026-03024-3.

55. Yurdal, M. O. & Kincal, R. Y. Effects of digital versus face-to-face peer support on clinical skills: a three-arm randomized trial. npj Digit. Med. 9, 683 (2026).

56. Welch, B. L. The generalization of ‘Student’s’ problem when several different population variances are involved. Biometrika 34, 28–35 (1947).

57. Cohen, J. Statistical Power Analysis for the Behavioral Sciences. (Lawrence Erlbaum Associates, Hillsdale, NJ, 1988).

58. Harris, C. R. et al. Array programming with NumPy. Nature 585, 357–362 (2020).

59. Virtanen, P. et al. SciPy 1.0: fundamental algorithms for scientific computing in Python. Nat. Methods 17, 261–272 (2020).

60. Seabold, S. & Perktold, J. Statsmodels: Econometric and Statistical Modeling with Python. in Proc. 9th Python in Science Conference 92–96 (SciPy, 2010). doi:10.25080/Majora-92bf1922-011.

61. Hunter, J. D. Matplotlib: A 2D Graphics Environment. Comput. Sci. Eng. 9, 90–95 (2007).

62. Liu, X., Cruz Rivera, S., Moher, D., Calvert, M. J. & Denniston, A. K. Reporting guidelines for clinical trial reports for interventions involving artificial intelligence: the CONSORT-AI extension. Nat. Med. 26, 1364–1374 (2020).

