## Supplementary material for "Calibrated large language models demonstrate a consistent advantage in a randomized longitudinal medical education intervention": Figures with legends, including Supplementary Figures S1 and S2

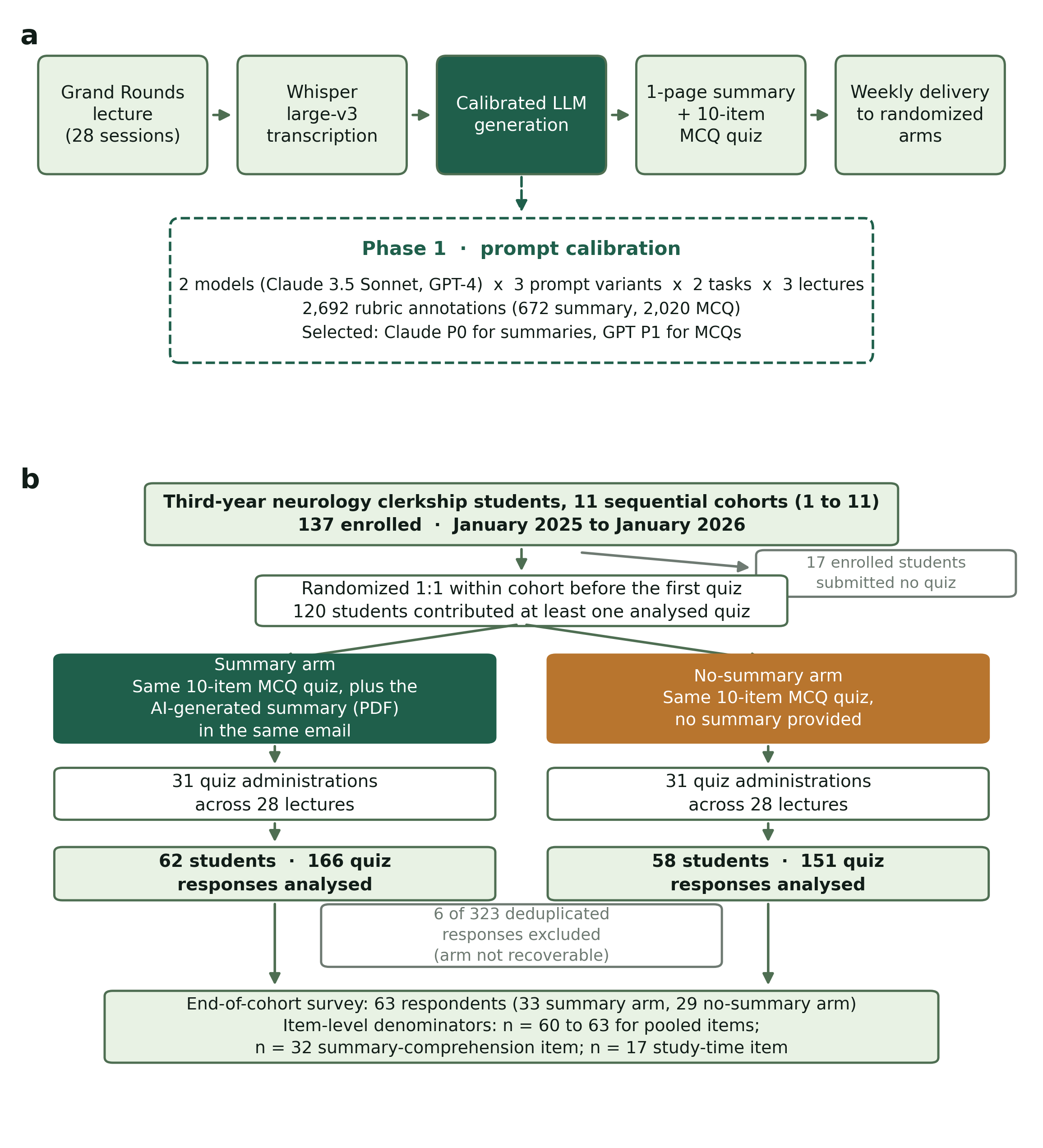

**Fig. 1 | Pipeline architecture and participant flow.** a, The four-stage generation pipeline. Recorded Grand Rounds audio is transcribed with Whisper large-v3, processed by a calibrated LLM under a structured prompt to produce a one-page summary and a ten-item MCQ quiz, and delivered to randomized arms. The dashed inset summarizes Phase 1 prompt calibration, in which two models and three prompt variants were scored across two tasks and three lectures using 2,692 rubric annotations, selecting Claude Prompt 0 for summaries and GPT Prompt 1 for MCQs. b, CONSORT-style flow for the Phase 2 randomized intervention. Of 137 students enrolled across 11 sequential cohorts, 17 submitted no quiz and 120 contributed at least one analyzed response (62 Summary, 58 No-summary), giving 317 analyzed responses from 323 collected.

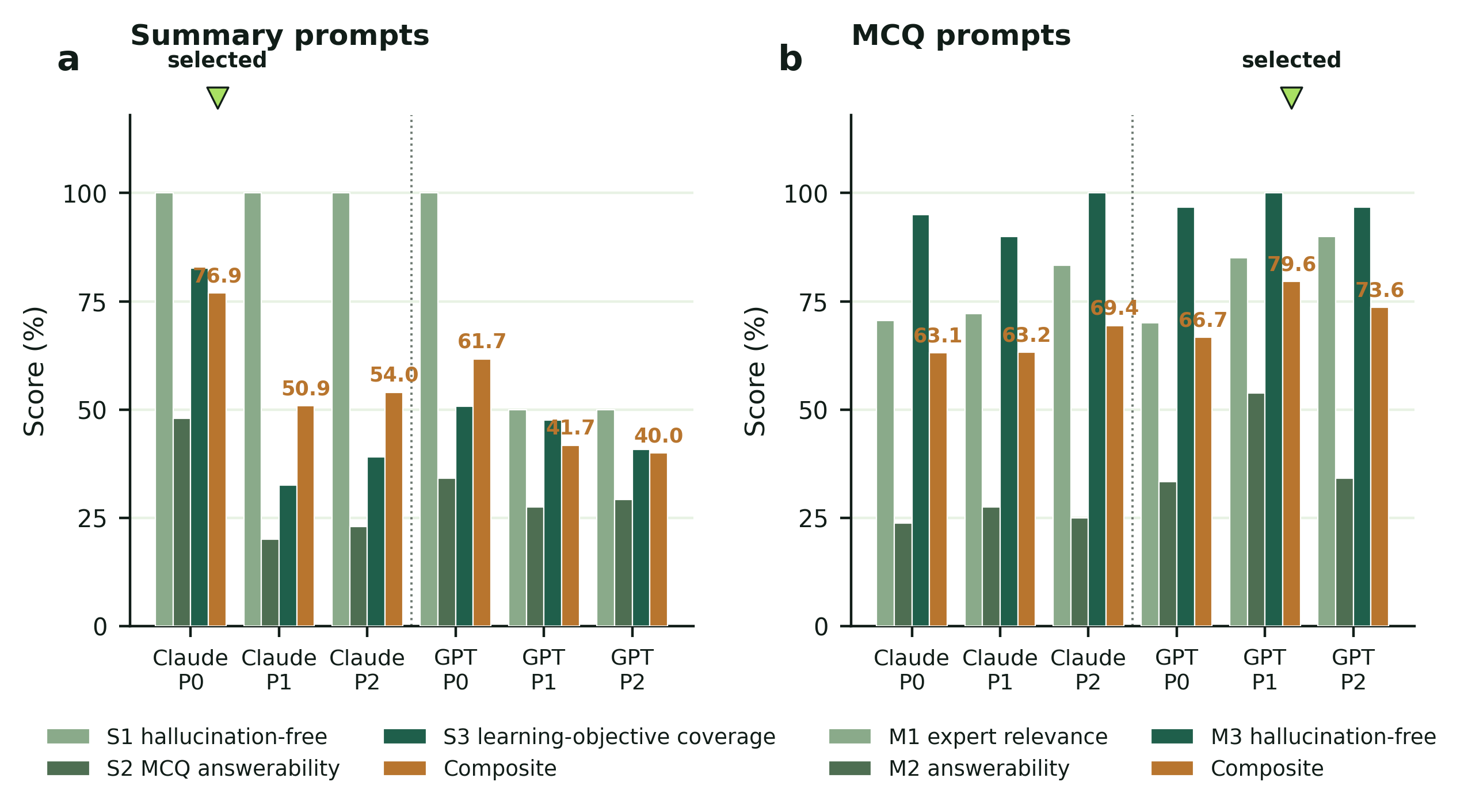

**Fig. 2 | Phase 1 prompt calibration composite scores.** a, Summary prompt variants scored on S1 (hallucination-free rate), S2 (overall MCQ answerability from the summary alone) and S3 (learning-objective coverage, the proportion of the lecture's stated learning objectives addressed by the summary, judged by trained reviewers), with the equal-weighted composite in amber. b, MCQ prompt variants scored on M1 (expert clinical relevance), M2 (answerability across candidate summaries) and M3 (MCQ hallucination-free rate). Green markers indicate the variant selected for deployment.

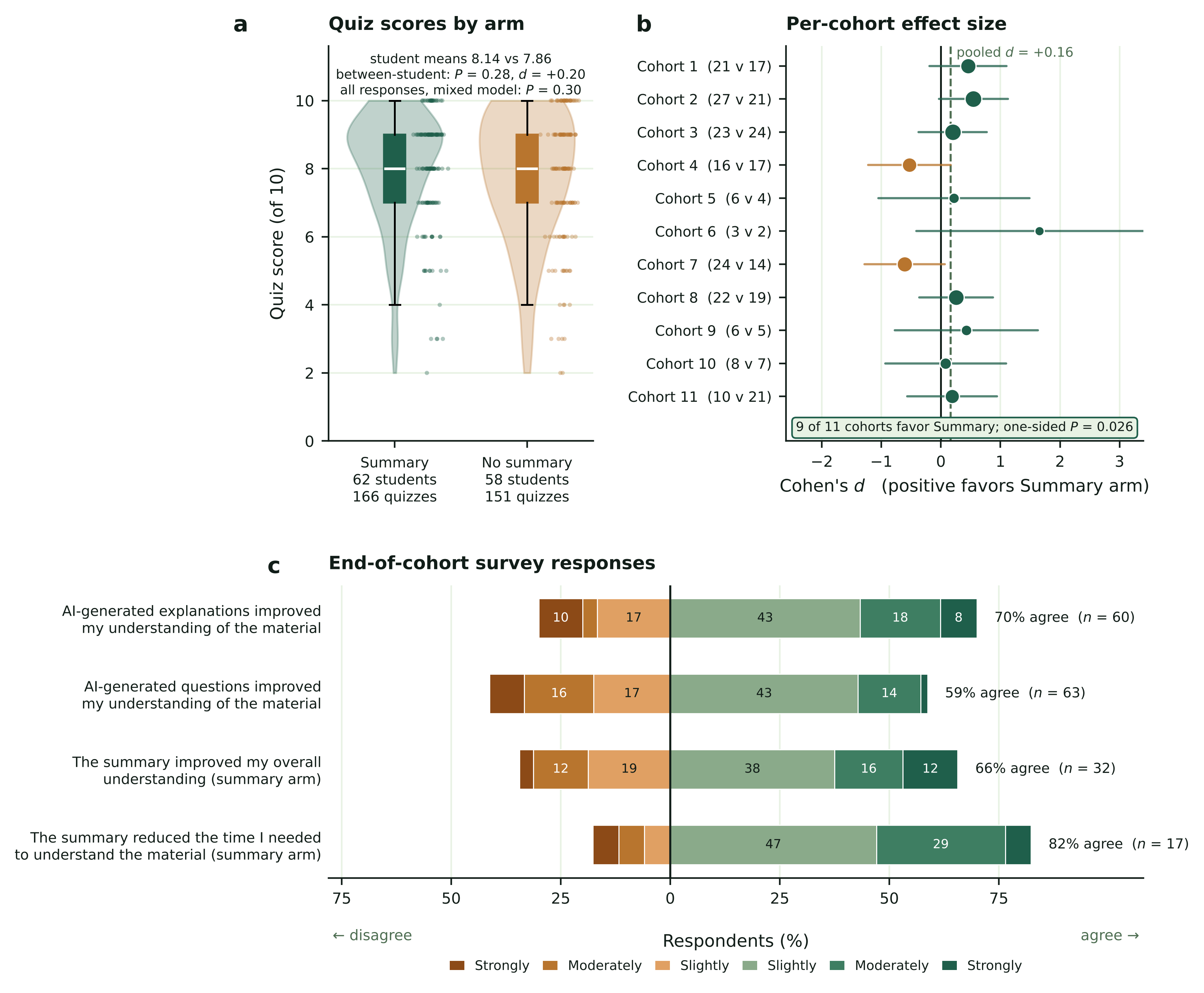

**Fig. 3 | Phase 2 intervention outcomes.** a, Distribution of individual quiz scores by arm (Annotations give the principal between-student comparison, in which each student contributes one mean score (Welch's t-test, P = 0.28; d = +0.20), and the mixed-effects model fitted to all responses (P = 0.30). b, Per-cohort standardized mean difference (Cohen's d) with approximate 95% confidence intervals; marker area is proportional to total responses, and per-cohort arm sizes are given in parentheses. Cohorts are numbered 1 to 11 in chronological order, and the dashed line marks the pooled response-level d (+0.16). The Summary arm scored higher in 9 of 11 cohorts (randomization test on the number of cohorts favoring Summary, permuting arm assignment among students within cohort, one-sided P = 0.026; exact binomial P = 0.033). c, End-of-cohort survey responses on a six-point Likert scale; denominators differ by item because summary-specific items were shown only to students who reported receiving a summary.

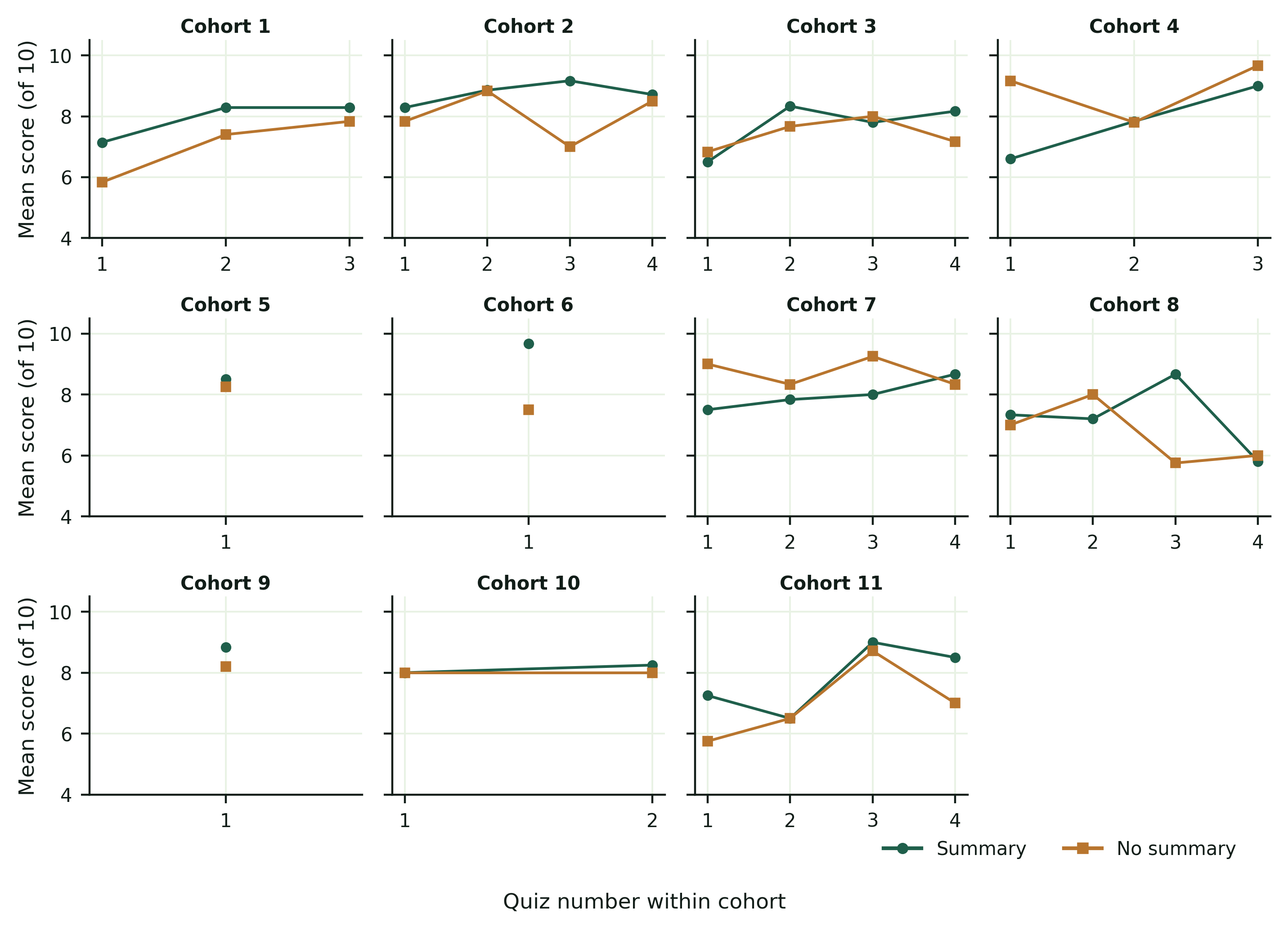

**Supplementary Fig. S1 | Per-quiz score trajectories by cohort.** Mean quiz score by arm for each quiz administration within each of the 11 cohorts, numbered 1 to 11 in chronological order. Green circles denote the Summary arm and amber squares the No-summary arm.

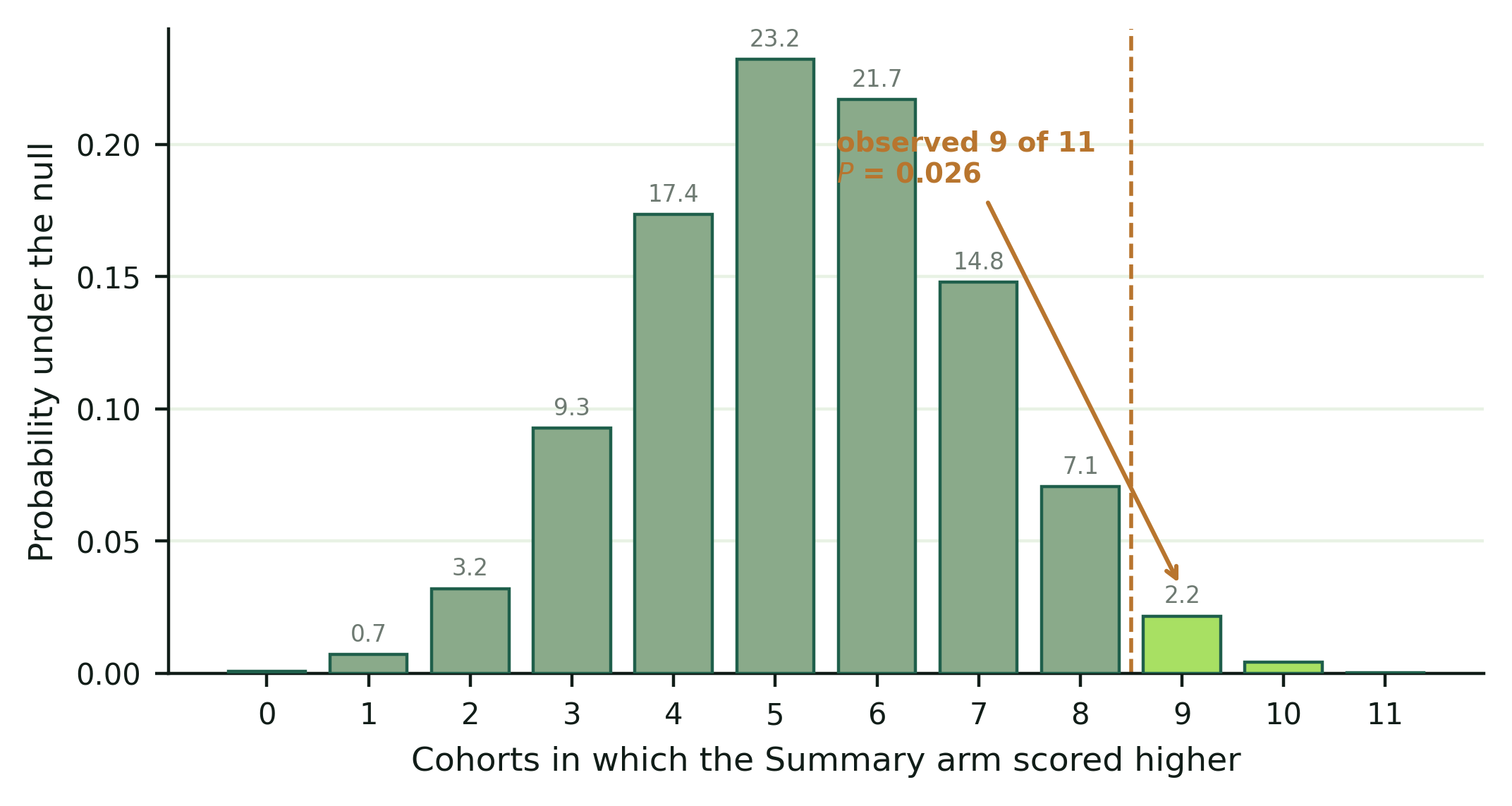

**Supplementary Fig. S2 | Randomization null distribution for the cohort-level result.** Distribution of the number of cohorts favoring the Summary arm under 100,000 permutations of arm assignment among students within cohort, which reproduces the actual randomization and preserves all within-student correlation. The null centers at 5.35 rather than 5.5 because allocation ratios differ across cohorts. The observed value of 9 of 11 falls in the upper tail (one-sided P = 0.026).
